# Elevated Rates of Gastrointestinal Dysfunction in Children with Neurodevelopmental Disabilities: Not Just an Autism Issue

**DOI:** 10.64898/2026.08.17.26360370

**Authors:** Juliann M. Savatt, Michelle Pistner Nixon, Alexander S.F. Berry, Alicia Johns, Lauren Kasparson Walsh, Christa Lese Martin, David H. Ledbetter, Thomas D. Challman, Scott M. Myers

## Abstract

Gastrointestinal (GI) conditions are common among children with neurodevelopmental disabilities (NDDs), and are associated with functional impairment, behavioral symptoms, and increased health care utilization. A unique relationship between autism and GI dysfunction has been proposed, leading to a focus on autism in GI research, management guidelines, and clinical tool development. Leveraging >20 years of electronic health record data and a cohort of 42,204 cases with attention-deficit/hyperactivity disorder, autism, cerebral palsy, epilepsy, or intellectual disability and 297,402 controls without NDDs, we quantified associations between NDDs and GI conditions in children. GI conditions were more common in cases than controls across all individual NDDs; intellectual disability and cerebral palsy were most strongly associated with having a GI condition. In this work, clinically recognized GI morbidity was elevated across all NDDs and not unique to autism, suggesting that a broader, transdiagnostic approach to GI dysfunction in children with NDDs is warranted.

## Introduction

Neurodevelopmental disabilities (NDDs), such as attention-deficit/hyperactivity disorder (ADHD), autism spectrum disorder (ASD), cerebral palsy (CP), epilepsy (EP), and intellectual disability (ID), affect approximately 17% of children in the United States^1,2^. In addition to their defining cognitive and/or behavioral features and functional impairments, NDDs are strongly associated with medical comorbidities that affect clinical presentation, prognosis, and management^3–12^. Gastrointestinal (GI) conditions are among the most common and impactful medical comorbidities observed in NDD populations^13–22^. Comorbid GI conditions frequently result in discomfort and impairment and are associated with emotional distress, behavior problems, psychiatric comorbidities, sleep disturbance, increased healthcare utilization, and reduced quality of life^17,21,23–28^.

The NDD-GI dysfunction relationship has been investigated most extensively in children with ASD, 33-49% of whom have GI conditions^14,15,29^. Overall (aggregate) GI problems and some specific conditions, including constipation, diarrhea, and abdominal pain, are significantly more prevalent in children with ASD than in typically developing controls, and these problems tend to persist throughout childhood^14,15,23,29,30^. However, no GI pathology specific to ASD has been identified^31,32^. Although they have garnered less general interest than the ASD-focused studies, elevated rates of aggregate GI problems and some specific GI conditions, such as constipation, have also been reported among individuals with ADHD, CP, EP, and ID relative to typically developing controls^13,16–20,22,33^.

In addition to the disproportionate focus on ASD, current understanding of the relationship between NDDs and GI dysfunction in the pediatric population is hampered by several other limitations in the existing literature, including failure to account for coexisting NDDs, methodological heterogeneity across studies, limited comparisons across NDD categories, and inconsistent findings regarding disorder-specific associations. Although it is well established that many individuals have more than one NDD^34,35^, published studies of NDD-GI relationships have not accounted for the possibility of coexisting NDDs. The absence of studies that investigate the relationships between NDDs and GI dysfunction across multiple NDDs simultaneously and account for co-occurring diagnoses has hindered identification of precise associations and determination of whether observations are unique to specific NDDs or shared across disorders. Further, methodological heterogeneity, including variability in the specific GI conditions studied, makes it difficult to compare results across studies. The small number of studies that have compared GI morbidity across NDD categories have largely compared individuals with ASD to those with one or more other NDDs, and the results are inconsistent; some have found significantly more GI problems in children with ASD^25,26,36–38^, whereas others have not identified a significant difference^24,39,40^.

Despite these methodological limitations and unresolved questions, the existence of a unique relationship between ASD and GI dysfunction has been proposed^41,42^, and ASD has been prioritized in GI-related care and research. Consensus recommendations for the evaluation and management of GI conditions in ASD have been published^43,44^, key research objectives have been prioritized^45^, and ASD-specific tools have been developed to measure GI symptoms^46,47^, while similar efforts have not been undertaken for other NDDs. Given the limitations of the literature, further studies are needed to understand whether this prioritization is warranted or whether broader strategies across NDDs are needed to help improve outcomes.

In this work, we provide the most comprehensive assessment of clinical GI morbidity across pediatric NDDs to date, in which we retrospectively examined >20 years of electronic health record (EHR) data from one healthcare system to explore associations between a range of clinically documented GI conditions and NDDs (ADHD, ASD, CP, EP, and ID) in a large pediatric cohort of >42,000 NDD cases and >297,000 controls with sufficient EHR data. The study methodology acknowledges that many children have more than one NDD and allows estimation of the independent association of each NDD with aggregate and common individual GI outcomes while accounting for co-occurring NDDs and demographic variables. This paper quantifies the associations between NDDs and GI conditions, individually and in aggregate.

## Methods

### Data Collection

Geisinger is a health system in Pennsylvania serving ∼1 million active patients. All patients born between 1989 and 2017 were eligible for inclusion. Data for eligible patients were collected from the EHR from December 1, 2002 (outpatient EHR implementation) or birth through the patient’s 18^th^ birthday or December 1, 2021, whichever was earlier. To ensure sufficient health history, analyses were restricted to patients who had ≥6 months between their first and last completed EHR encounters during the study period. The study protocol was submitted to the Geisinger IRB review and did not meet the definition of human subject research as defined in 45 CFR 46.102(f).

Demographic information, including age, sex, ethnicity, and race, were extracted for each patient. Eight patients had unknown sex and were excluded from all analyses. NDDs, GI conditions, and feeding difficulties were defined via relevant diagnosis codes (eTable 1). All inpatient, outpatient, and telemedicine encounters and problem list entries using these codes were extracted for each patient during the study period.

### Defining Cases and Controls

For both NDDs and GI conditions, a patient was considered to have that disorder if a relevant code was used in ≥2 outpatient or telemedicine visits, ≥1 inpatient visit, or a problem list entry. For GI conditions, emergency visits were additionally considered when determining presence of the diagnosis along with outpatient and telemedicine visits (i.e., a patient with ≥2 outpatient, telemedicine visits, or emergency visits was considered to have the condition). Patients were also considered to be NDD cases if they had one or more outpatient or telemedicine visits with a developmental medicine, pediatric neurology, or pediatric psychology or psychiatry department clinician where an NDD diagnosis code was used.

Cases included in further analyses were defined as those meeting criteria for one of five NDDs: ADHD, ASD, CP, EP, and ID. Patients classified as only having specific learning disorders, Diagnostic and Statistical Manual of Mental Disorders, Fifth Edition (DSM-5) motor disorders^48^, communication disorders, or other/non-specific neurodevelopmental disorders (eTable 1) were excluded from further analyses. These diagnoses were excluded due to concerns that they were inconsistently captured in the EHR. Any child not meeting the above criteria for any defined NDD was considered a control.

### Statistical Analysis

A basic descriptive analysis was conducted to compare demographics and the proportion of GI disorders between those who had any of the five NDDs of interest (ADHD, ASD, CP, EP, and ID; termed “any NDD”) and controls who had no reported NDDs. All cases and controls were assessed for 14 specific GI conditions and feeding difficulties as described (eTable 1). For analyses, two separate aggregate variables were created 1) having any of the 14 GI diagnoses, termed “any GI condition” and 2) having any of the 14 GI diagnoses or feeding difficulties, termed “any GI condition or feeding difficulty.” Furthermore, patients who were considered cases were sub-stratified by their number of NDDs. For both analyses, categorical variables were summarized by counts and percentages, and continuous variables were summarized using median and interquartile range (IQR). Comparisons were conducted between groups. For categorical variables, Pearson’s Chi-square test, and, if assumptions were unmet, Fisher’s exact test were used. For continuous variables, the Wilcoxon rank sum test was employed.

To adjust for any differences in NDD and GI diagnosis proportions attributable to sex, age, race, or length of EHR during the study period, cases and controls were matched in a 2:1 ratio. To match cases to controls, sex and race were matched exactly, age relative to January 1, 1989, and age at last inpatient, outpatient, or telemedicine encounter were matched within three years, and the length of EHR was matched to within one year. Love plots and standardized mean differences were assessed to ensure balance^49^. In an analogous manner to the analyses of unmatched data, comparisons between the proportions of GI conditions were performed on the matched cohort.

Because demographic variables and coexisting NDDs may also be associated with specific GI conditions, Firth’s bias corrected logistic regression was used to model the presence of specific GI conditions on the matched cohort for any GI condition that occurred in >5% of case patients. This included the presence of any GI condition, constipation, GERD, nausea or vomiting, abdominal pain, and diarrhea. All models were fit using all available matched patients, and all models used the same set of explanatory variables: all five NDDs, all two-way interactions between these conditions, sex, race, age at study start, age at last encounter during the study period, and length of EHR during the study period. To ensure all model coefficients had meaningful interpretation at zero, age at study start, age at last encounter, and length of EHR were mean centered. Matching weights were used for all analyses on matched data. Standard model diagnostics were used to assess model fit.

Within each model, all one-way coefficients for each NDD were statistically compared with all other one-way coefficients for the remaining NDDs, resulting in 10 comparisons within each model using Chi-square tests as implemented by the *linearHypothesis* function in the *car* package^50^.

All analyses were conducted in R, version 4.4.1^51^. Matching was conducted using the *MatchIt* package, version 4.7.0^52^. Firth’s bias corrected logistic regression was fit using the *brglm* package, version 0.7.3^53^. For all tests, a Bonferroni-adjusted α of 0.0001 was used^54^. The reported 95% confidence intervals were unadjusted.

## Results

After extracting EHR data and applying casing decisions as described, 42,204 patients (12.4%) were identified as cases for one of the five studied NDDs, and 297,402 patients were identified as NDD-negative controls. Most cases only had a single NDD (N=33,194; 78.65%; eTable 2) and rates of the five NDDs were similar to those summarized in relevant prevalence surveys^1,2^ (eTable 3). Patients with multiple NDDs were, on average, younger at last encounter, more likely to be male, and more likely to have a GI condition relative to those with a single NDD (eTable 4). Figure 1 includes a description of the study pipeline and cohort characteristics.

**Figure 1.**
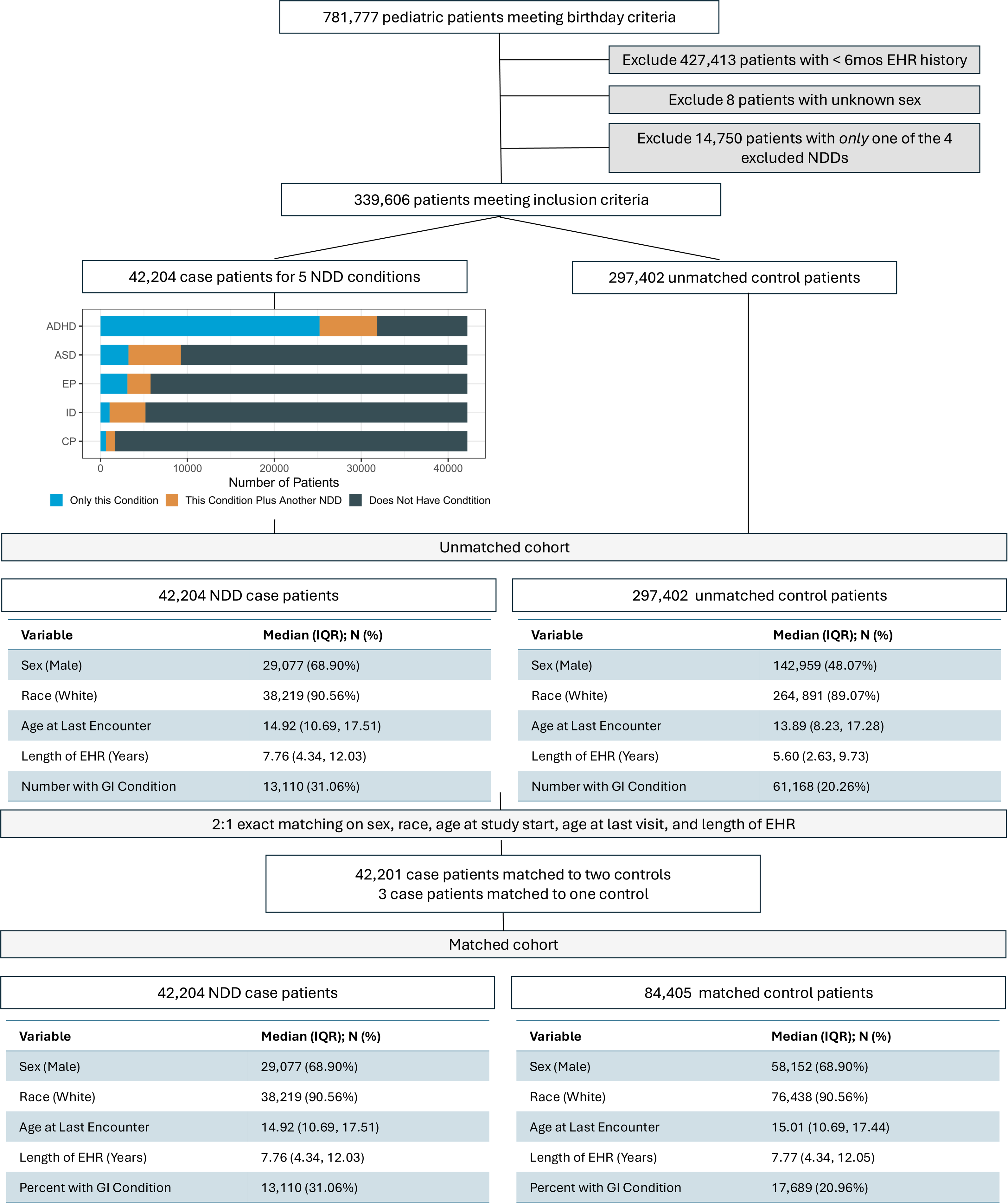
Description of the Study Pipeline, Cohort Characteristics, and Demographics. NDD, Neurodevelopmental disability; GI – Gastrointestinal. The NDD diagnoses of interest included ADHD - attention-deficit/hyperactivity disorder, ASD - autism spectrum disorder, CP - cerebral palsy, EP - epilepsy, ID - intellectual disabilities.

Demographic comparisons between cases and unmatched controls showed cases were more likely to be male (mean difference: 20.83%; p-value <0.0001) and marginally older at last encounter (mean difference: 0.47 years; p-value <0.0001) (eTables 5 and 6; Figure 1). To minimize the effects of these differences, matching was performed. Except for three cases who matched to one control, all cases matched to two controls. Matching greatly improved balance across selected covariates (standardized mean difference: 0.0002); no significant differences between sex, race, age at the start of the study period, age at last encounter during the study period, and length of EHR were observed (Figure 1; eTable 7).

The frequency of having any GI diagnosis was higher in NDD cases (31.06%) compared to matched controls (22.29%) (mean difference: 8.77%; p-value <0.0001). Feeding difficulties and 10 of 14 individual GI conditions studied were significantly more common in cases relative to controls (Table 1; eTable 8). These results largely held when studying individual NDDs in analyses that included all relevant cases; all individual NDD diagnoses were associated with a higher proportion of aggregate GI conditions when compared to matched controls (Table 1; range of mean differences: 8.00% (ADHD, p<0.0001) to 27.19% (CP, p<0.0001). There were many associations between individual NDDs and specific GI conditions. For example, eosinophilic esophagitis was significantly more common relative to controls in cases with either CP or ID, but not in cases with ASD, ADHD, or EP.

**Table 1.** Rates of GI Conditions by Matched Cases versus Controls.

|  | Any NDD |  |  | ADHD |  |  | ASD |  |  | CP |  |  | EP |  |  | ID |  |  |
| --- | --- | --- | --- | --- | --- | --- | --- | --- | --- | --- | --- | --- | --- | --- | --- | --- | --- | --- |
|  | Case<br>N =<br>42,204<br>n (%) | Control<br>N =<br>84,405<br>n (%) | p-<br>val<br>ue | Case<br>N =<br>31,832<br>n (%) | Control<br>N =<br>63,662<br>n (%) | p-<br>val<br>ue | Case<br>N =<br>9,241<br>n (%) | Contr<br>ol N =<br>18,481<br>n (%) | p-<br>val<br>ue | Case<br>N = 1,645<br>n (%) | Control<br>N =<br>3,289<br>n (%) | p-<br>val<br>ue | Case<br>N = 5,761<br>n (%) | Control<br>N =<br>11,522<br>n (%) | p-<br>val<br>ue | Case<br>N = 5,173<br>n (%) | Contr<br>ol N =<br>10,344<br>n (%) | p-<br>val<br>ue |
| Any GI condition | 13,110<br>(31.06<br>%) | 18,816<br>(22.29<br>%) | <0.<br>000<br>1 | 9,699<br>(30.47%) | 14,302<br>(22.47%) | <0.<br>00<br>01 | 2,861<br>(30.96<br>%) | 4,034<br>(21.83<br>%) | <0.<br>00<br>01 | 815<br>(49.54%) | 735<br>(22.35<br>%) | <0.<br>00<br>01 | 2,030<br>(35.24%) | 2,684<br>(23.29%<br>) | <0.<br>00<br>01 | 2,144<br>(41.45%) | 2,456<br>(23.74<br>%) | <0.<br>00<br>01 |
| Feeding difficulties | 1,855<br>(4.40%) | 662<br>(0.78%) | <0.<br>000<br>1 | 718<br>(2.26%) | 476<br>(0.75%) | <0.<br>00<br>01 | 627<br>(6.78%) | 141<br>(0.76<br>%) | <0.<br>00<br>01 | 451<br>(27.42%) | 35<br>(1.06%) | <0.<br>00<br>01 | 611<br>(10.61%) | 96<br>(0.83%) | <0.<br>00<br>01 | 988<br>(19.10%) | 124<br>(1.20<br>%) | <0.<br>00<br>01 |
| Any GI condition or Feeding difficulties | 13,553<br>(32.11<br>%) | 19,052<br>(22.57<br>%) | <0.<br>000<br>1 | 9,907<br>(31.12%) | 14,467<br>(22.72%) | <0.<br>00<br>01 | 3,072<br>(33.24<br>%) | 4,088<br>(22.12<br>%) | <0.<br>00<br>01 | 859<br>(52.22%) | 745<br>(22.65<br>%) | <0.<br>00<br>01 | 2,103<br>(36.50%) | 2,720<br>(23.61%<br>) | <0.<br>00<br>01 | 2,334<br>(45.12%) | 2,502<br>(24.19<br>%) | <0.<br>00<br>01 |
| Constipation | 5,415<br>(12.83<br>%) | 5,381<br>(6.38%) | <0.<br>000<br>1 | 3,756<br>(11.80%) | 4,057<br>(6.37%) | <0.<br>00<br>01 | 1,441<br>(15.59<br>%) | 1,194<br>(6.46<br>%) | <0.<br>00<br>01 | 502<br>(30.52%) | 211<br>(6.42%) | <0.<br>00<br>01 | 972<br>(16.87%) | 758<br>(6.58%) | <0.<br>00<br>01 | 1,237<br>(23.91%) | 730<br>(7.06<br>%) | <0.<br>00<br>01 |
| Nausea or vomiting | 4,286<br>(10.16<br>%) | 5,472<br>(6.48%) | <0.<br>000<br>1 | 3,203<br>(10.06%) | 4,230<br>(6.64%) | <0.<br>00<br>01 | 884<br>(9.57%) | 1,167<br>(6.31<br>%) | <0.<br>00<br>01 | 235<br>(14.29%) | 214<br>(6.51%) | <0.<br>00<br>01 | 764<br>(13.26%) | 795<br>(6.90%) | <0.<br>00<br>01 | 726<br>(14.03%) | 722<br>(6.98<br>%) | <0.<br>00<br>01 |
| Gastroesophageal reflux disease | 5,200<br>(12.32<br>%) | 6,343<br>(7.51%) | <0.<br>000<br>1 | 3,625<br>(11.39%) | 4,817<br>(7.57%) | <0.<br>00<br>01 | 1,049<br>(11.35%<br>) | 1,323<br>(7.16<br>%) | <0.<br>00<br>01 | 545<br>(33.13%) | 256<br>(7.78%) | <0.<br>00<br>01 | 1,003<br>(17.41%) | 880<br>(7.64%) | <0.<br>00<br>01 | 1,099<br>(21.24%) | 813<br>(7.86<br>%) | <0.<br>00<br>01 |
| Abdominal pain or tenderness | 3,876<br>(9.18%) | 6,645<br>(7.87%) | <0.<br>000<br>1 | 3,119<br>(9.80%) | 5,173<br>(8.13%) | <0.<br>00<br>01 | 686<br>(7.42%) | 1,293<br>(7.00<br>%) | 0.1<br>9 | 146<br>(8.88%) | 256<br>(7.78%) | 0.1<br>9 | 549<br>(9.53%) | 942<br>(8.18%) | 0.0<br>03 | 392 (7.58%) | 734<br>(7.10<br>%) | 0.2<br>8 |
| Diarrhea | 2,875<br>(6.81%) | 3,667<br>(4.34%) | <0.<br>000<br>1 | 2,154<br>(6.77%) | 2,845<br>(4.47%) | <0.<br>00<br>01 | 645<br>(6.98%) | 753<br>(4.07<br>%) | <0.<br>00<br>01 | 153<br>(9.30%) | 133<br>(4.04%) | <0.<br>00<br>01 | 440<br>(7.64%) | 482<br>(4.18%) | <0.<br>00<br>01 | 537<br>(10.38%) | 475<br>(4.59<br>%) | <0.<br>00<br>01 |
| Functional dyspepsia (indigestion) or heartburn | 863<br>(2.04%) | 1,391<br>(1.65%) | <0.<br>000<br>1 | 686<br>(2.16%) | 1,105<br>(1.74%) | <0.<br>00<br>01 | 131<br>(1.42%) | 247<br>(1.34<br>%) | 0.5<br>8 | 50 (3.04%) | 57<br>(1.73%) | 0.0<br>03 | 141<br>(2.45%) | 224<br>(1.94%) | 0.0<br>3 | 101 (1.95%) | 135<br>(1.31<br>%) | 0.0<br>02 |
| Flatulence and related conditions | 271<br>(0.64%) | 308<br>(0.36%) | <0.<br>000<br>1 | 158<br>(0.50%) | 236<br>(0.37%) | 0.0<br>04 | 85<br>(0.92%) | 67<br>(0.36<br>%) | <0.<br>00<br>01 | 40 (2.43%) | 7<br>(0.21%) | <0.<br>00<br>01 | 67<br>(1.16%) | 47<br>(0.41%) | <0.<br>00<br>01 | 101 (1.95%) | 38<br>(0.37<br>%) | <0.<br>00<br>01 |
| Irritable Bowel Syndrome | 264<br>(0.63%) | 495<br>(0.59%) | 0.4 | 213<br>(0.67%) | 398<br>(0.63%) | 0.4<br>2 | 55<br>(0.60%) | 77<br>(0.42<br>%) | 0.0<br>4 | 4 (0.24%) | 10<br>(0.30%) | 0.7<br>9 | 27<br>(0.47%) | 70<br>(0.61%) | 0.2<br>5 | 13 (0.25%) | 44<br>(0.43<br>%) | 0.0<br>9 |
| Eosinophilic esophagitis | 151<br>(0.36%) | 237<br>(0.28%) | 0.0<br>2 | 92<br>(0.29%) | 187<br>(0.29%) | 0.9 | 40<br>(0.43%) | 53<br>(0.29 | 0.0<br>5 | 27 (1.64%) | 10<br>(0.30%) | <0.<br>00 | 32<br>(0.56%) | 24<br>(0.21%) | 2e-04 | 45 (0.87%) | 22<br>(0.21 | <0.<br>00 |

|  |  |  |  |  |  |  |  | %) |  |  |  | **01** |  |  |  |  | %) | **01** |
| --- | --- | --- | --- | --- | --- | --- | --- | --- | --- | --- | --- | --- | --- | --- | --- | --- | --- | --- |
| Gastroparesis | 199<br>(0.47%) | 124<br>(0.15%) | **<0.0001** | 98<br>(0.31%) | 87<br>(0.14%) | **<0.0001** | 33<br>(0.36%) | 31<br>(0.17%) | 0.002 | 34 (2.07%) | 6<br>(0.18%) | **<0.0001** | 69<br>(1.20%) | 18<br>(0.16%) | **<0.0001** | 87 (1.68%) | 16<br>(0.15%) | **<0.0001** |
| Crohn's Disease | 85<br>(0.20%) | 196<br>(0.23%) | 0.27 | 60<br>(0.19%) | 163<br>(0.26%) | 0.04 | 21<br>(0.23%) | 33<br>(0.18%) | 0.39 | 4 (0.24%) | 6<br>(0.18%) | 0.74 | 12<br>(0.21%) | 22<br>(0.19%) | 0.81 | 13 (0.25%) | 17<br>(0.16%) | 0.25 |
| Ulcerative colitis | 48<br>(0.11%) | 85<br>(0.10%) | 0.5 | 26<br>(0.08%) | 70<br>(0.11%) | 0.19 | 17<br>(0.18%) | 17<br>(0.09%) | 0.04 | 7 (0.43%) | 0<br>(0.00%) | 5e-04 | 13<br>(0.23%) | 7<br>(0.06%) | 0.03 | 14 (0.27%) | 7<br>(0.07%) | 0.01 |
| Other noninfective colitis | 1,041<br>(2.47%) | 1,356<br>(1.61%) | **<0.0001** | 756<br>(2.37%) | 1,025<br>(1.61%) | **<0.0001** | 244<br>(2.64%) | 300<br>(1.62%) | **<0.0001** | 53 (3.22%) | 58<br>(1.76%) | 0.001 | 161<br>(2.79%) | 199<br>(1.73%) | **<0.0001** | 210 (4.06%) | 199<br>(1.92%) | **<0.0001** |
| Intestinal malabsorption | 393<br>(0.93%) | 420<br>(0.50%) | **<0.0001** | 227<br>(0.71%) | 287<br>(0.45%) | **<0.0001** | 113<br>(1.22%) | 108<br>(0.58%) | **<0.0001** | 21 (1.28%) | 17<br>(0.52%) | 0.004 | 75<br>(1.30%) | 67<br>(0.58%) | **<0.0001** | 127 (2.46%) | 83<br>(0.80%) | **<0.0001** |

Abbreviations: ADHD, attention-deficit/hyperactivity disorder; ASD, autism spectrum disorder; CP, cerebral palsy; EP, epilepsy; GI, gastrointestinal; ID, Intellectual disabilities; NDD, neurodevelopmental disability
<sup>a</sup>For all tests, a Bonferroni-adjusted $\alpha$ of 0.0001 was used. Significant p-values under this adjustment are bold.

Initial comparisons were performed separately for each NDD allowing individuals with multiple NDDs to contribute to more than one group. To account for overlap among co-occurring NDDs, Firth’s bias-corrected logistic regression models included all five NDDs simultaneously, allowing the association of each individual NDD with GI conditions to be estimated while adjusting for the others. Models were fit for any GI condition (the aggregate) and, separately, for the five individual GI conditions occurring in >5% of cases (constipation, GERD, nausea or vomiting, abdominal pain, and diarrhea). While all model coefficients are reported in Table 2, one-way coefficients from these models for each of the five NDDs are shown in Figure 2. By design, the odds ratios (OR) for all one-way interactions denote the effect of a given NDD on the GI condition for a white, male patient of average age with no other NDD. When evaluating any GI condition (the aggregate), all estimated odds ratios for the five NDD diagnoses were statistically greater than 1 (minimum OR [EP]: 1.28, 95% CI: 1.18, 1.39; maximum OR [ID]: 2.16, 95% CI: 1.94, 2.42). Across individual GI conditions, estimated ORs for each NDD were generally >1. Most of these were significant at the adjusted α with several exceptions for ASD, CP, ID, and EP (Figure 2). Otherwise, these models indicated that all NDDs are associated with at least 28% higher odds of any GI condition, including higher odds of constipation (37%). All NDDs, except for ASD, are associated with at least 49% higher odds of GERD. The other specific GI conditions examined (nausea/vomiting, abdominal pain, and diarrhea) were more variable in their association with individual NDDs (Figure 2).

**Figure 2.**
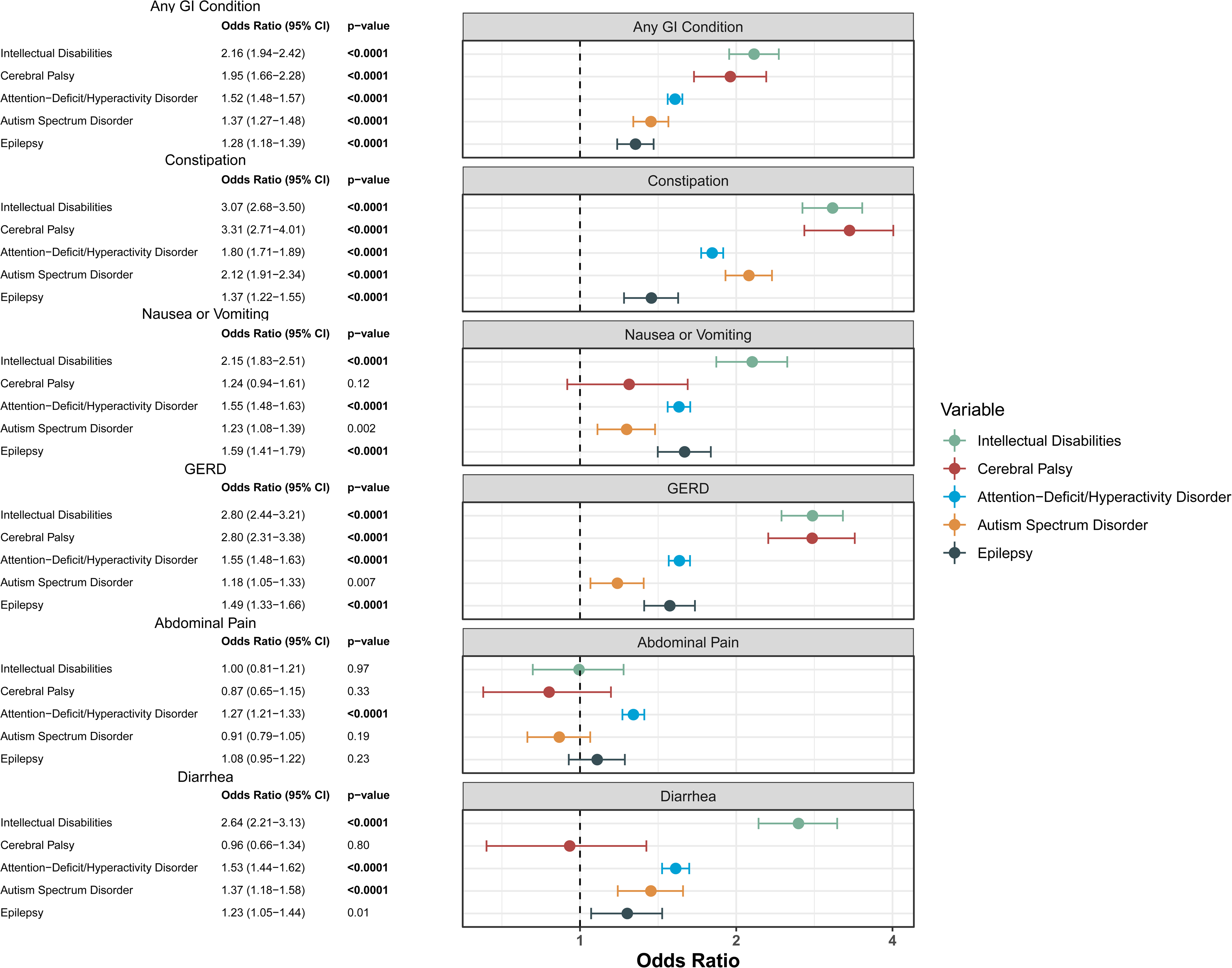
Estimated Odds Ratios for the NDD Main Effects for Selected GI Conditions. For all tests, a Bonferroni-adjusted α of 0.0001 was used. Bolded p-values indicate Bonferroni-adjusted significance. The reported confidence intervals were unadjusted 95% confidence intervals. Note that although several unadjusted 95% CIs do not cross 1 (cerebral palsy and nausea or vomiting, autism spectrum disorder and GERD, intellectual disability and abdominal pain, autism spectrum disorder and abdominal pain), they are not significant using the Bonferroni-adjusted α of 0.0001. GI, Gastrointestinal; GERD, gastroesophageal reflux disease.

**Table 2.**
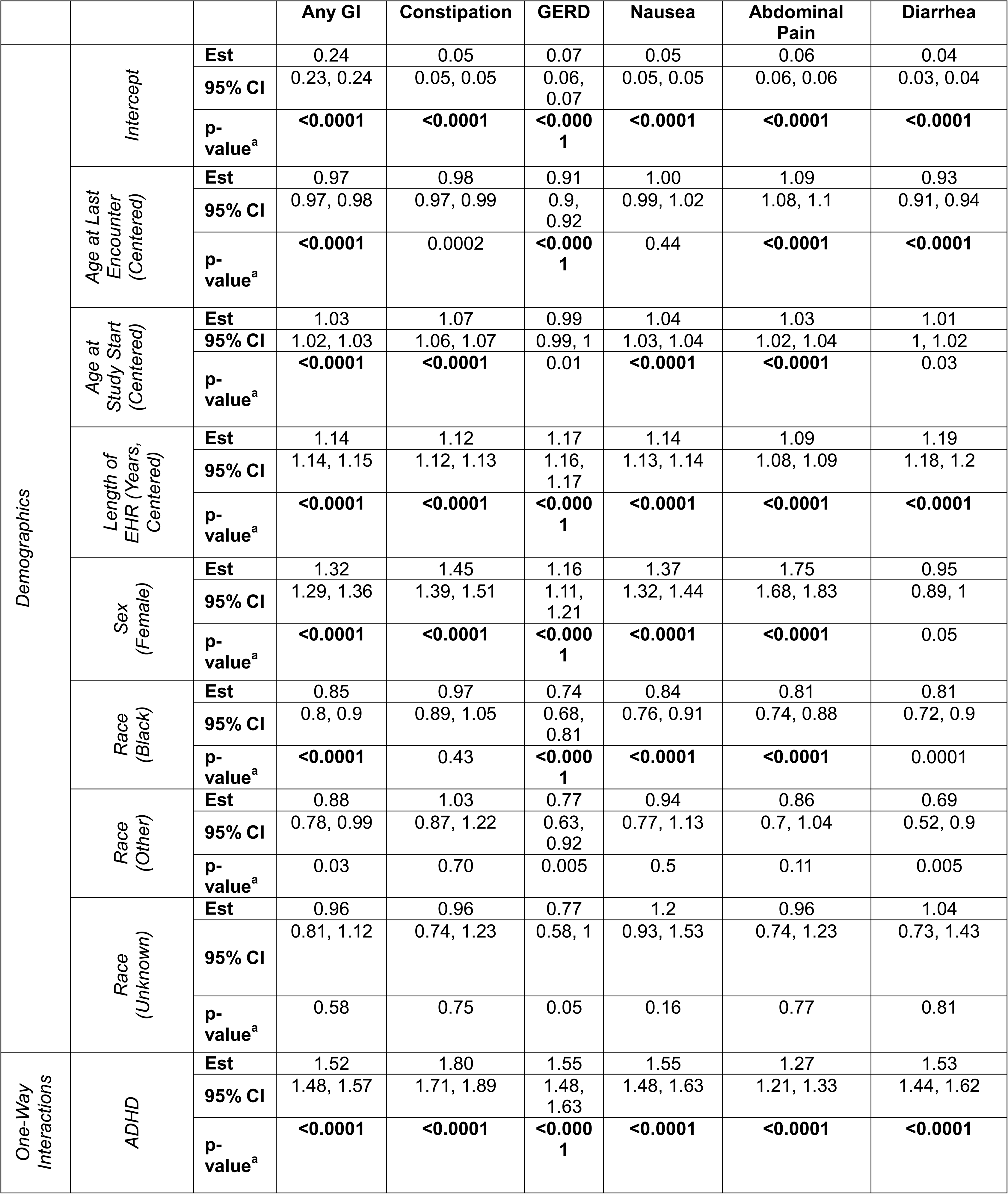
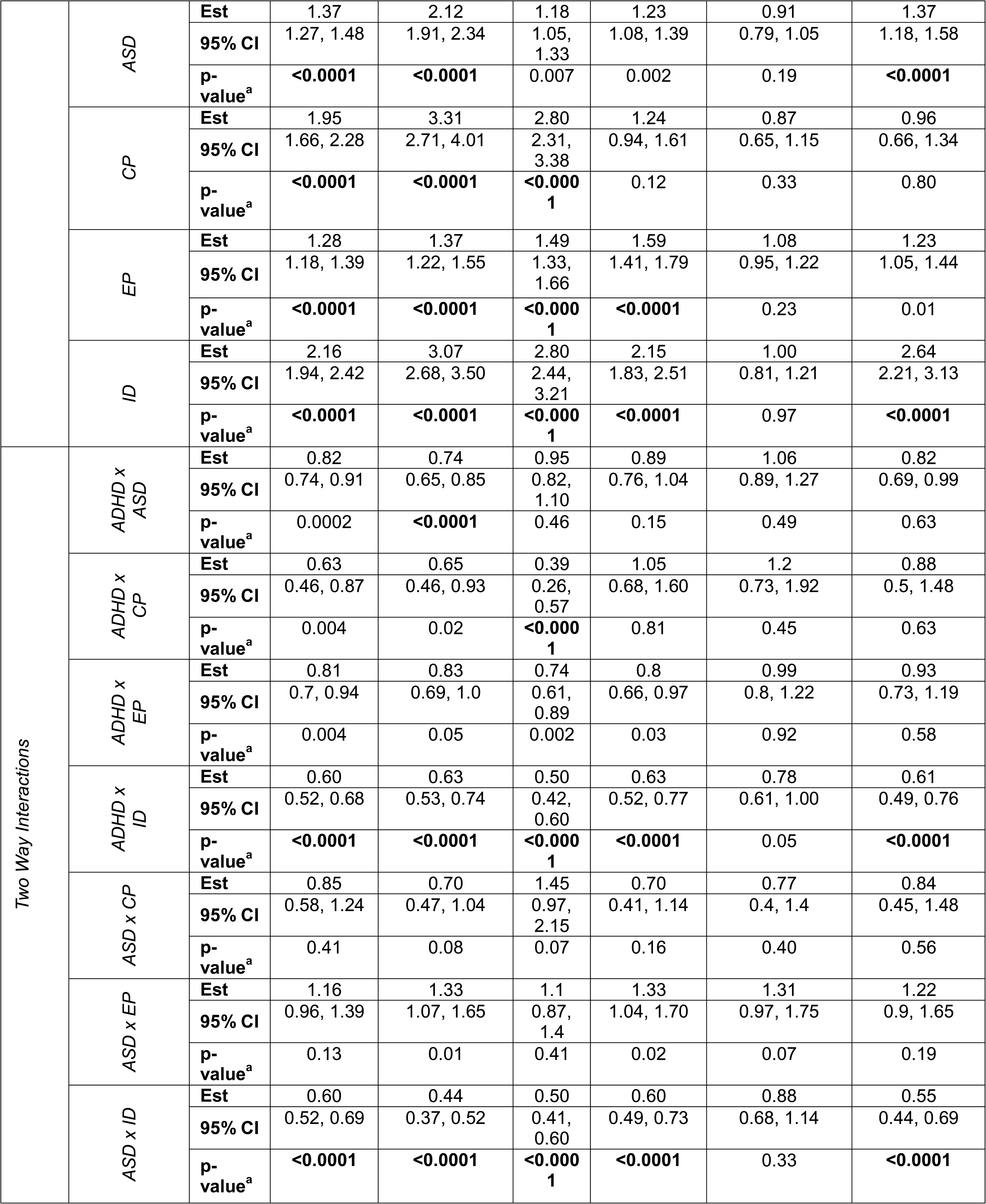
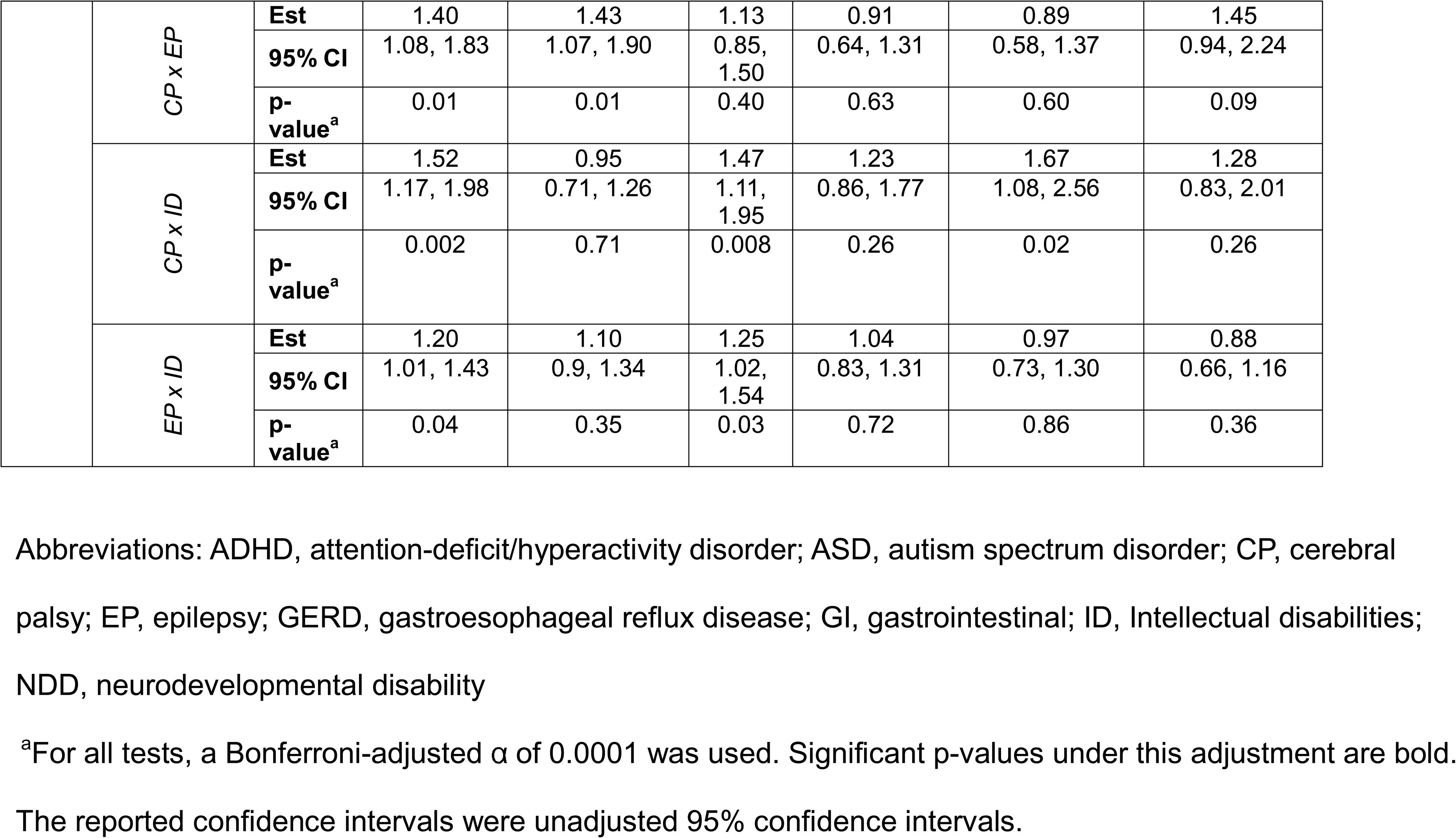
Logistic Regression Coefficients for GI Models.

When comparing one-way coefficients for any GI condition (the aggregate) for each NDD to the remaining NDDs, ID and CP had similar coefficients to one another (ID OR: 2.16 vs CP OR: 1.95, p=0.30) and larger coefficients than ASD (ID OR 2.16 vs ASD OR 1.37, p<0.0001, CP OR 1.95 vs ASD OR 1.37, p<0.0001) and EP (ID OR 2.16 vs EP OR 1.28, p<0.0001, CP OR 1.95 vs EP OR 1.28, p<0.0001); ID also had a higher coefficient than ADHD (ID OR 2.16 vs ADHD OR 1.52, p<0.0001) (Table 4). ADHD had higher a coefficient than EP (ADHD OR 1.52 vs EP OR 1.28 p<0.0001). No differences were observed between ADHD and CP (ADHD OR 1.52 vs CP OR 1.95, p=0.003), ADHD and ASD (ADHD OR 1.52 vs ASD OR 1.37, p=0.011), or ASD and EP (ASD OR 1.37 vs EP OR 1.28, p=0.23). When examining the five specific GI conditions (constipation, nausea/vomiting, GERD, abdominal pain, and diarrhea), patterns varied in terms of whether one-way coefficients among individual NDDs differed significantly (Table 4). ID and CP were most strongly associated with constipation and GERD, and ID was most strongly associated with diarrhea (Table 4).

To further study the association between GI conditions and NDDs, a sensitivity analysis was performed restricted to cases with record of only a single NDD versus their matched controls (Table 3; eTable 9). In this sub-analysis, higher frequencies of GI diagnoses were observed for all NDDs compared to matched controls (mean difference: 7.48%; p-value <0.0001) with the highest observed difference between those with ID and matched controls (mean difference: 18.18%; p-value <0.0001). With some exceptions, the frequency of GI conditions was higher in cases with single NDD compared to matched controls (eTable 9).

**Table 3.** Frequency of Various GI Conditions by NDD for Cases with Only One NDD Diagnosis.

|  | Any GI Condition |  |  | Any GI Condition or Feeding Difficulty |  |  |
| --- | --- | --- | --- | --- | --- | --- |
|  | Any GI Condition | Any GI Condition in Matched Controls | p-value<sup>a</sup> | Any GI Condition or Feeding Difficulty | Any GI Condition or Feeding Difficulty in Matched Controls | p-value |
| Any NDD (N = 33,194) | 9,826 (29.60%) | 14,682 (22.12%) | **<0.0001** | 10,064 (30.32%) | 14,857 (22.38%) | **<0.0001** |
| ADHD (N = 25,200) | 7,510 (29.80%) | 11,282 (22.39%) | **<0.0001** | 7,606 (30.18%) | 11,412 (22.64%) | **<0.0001** |
| ASD (N = 3,219) | 887 (27.56%) | 1,336 (20.75%) | **<0.0001** | 957 (29.73%) | 1,355 (21.05%) | **<0.0001** |
| CP (N = 633) | 202 (31.91%) | 244 (19.27%) | **<0.0001** | 216 (34.12%) | 248 (19.59%) | **<0.0001** |
| EP (N = 3,097) | 797 (25.73%) | 1,340 (21.63%) | **<0.0001** | 814 (26.28%) | 1,355 (21.88%) | **<0.0001** |
| ID (N = 1,045) | 430 (41.15%) | 480 (22.97%) | **<0.0001** | 471 (45.07%) | 487 (23.30%) | **<0.0001** |

Abbreviations: ADHD, attention-deficit/hyperactivity disorder; ASD, autism spectrum disorder; CP, cerebral palsy; EP, epilepsy; GI, gastrointestinal; ID, Intellectual disabilities; NDD, neurodevelopmental disability
<sup>a</sup>For all tests, a Bonferroni-adjusted $\alpha$ of 0.0001 was used. Significant p-values under this adjustment are bold.

**Table 4.**
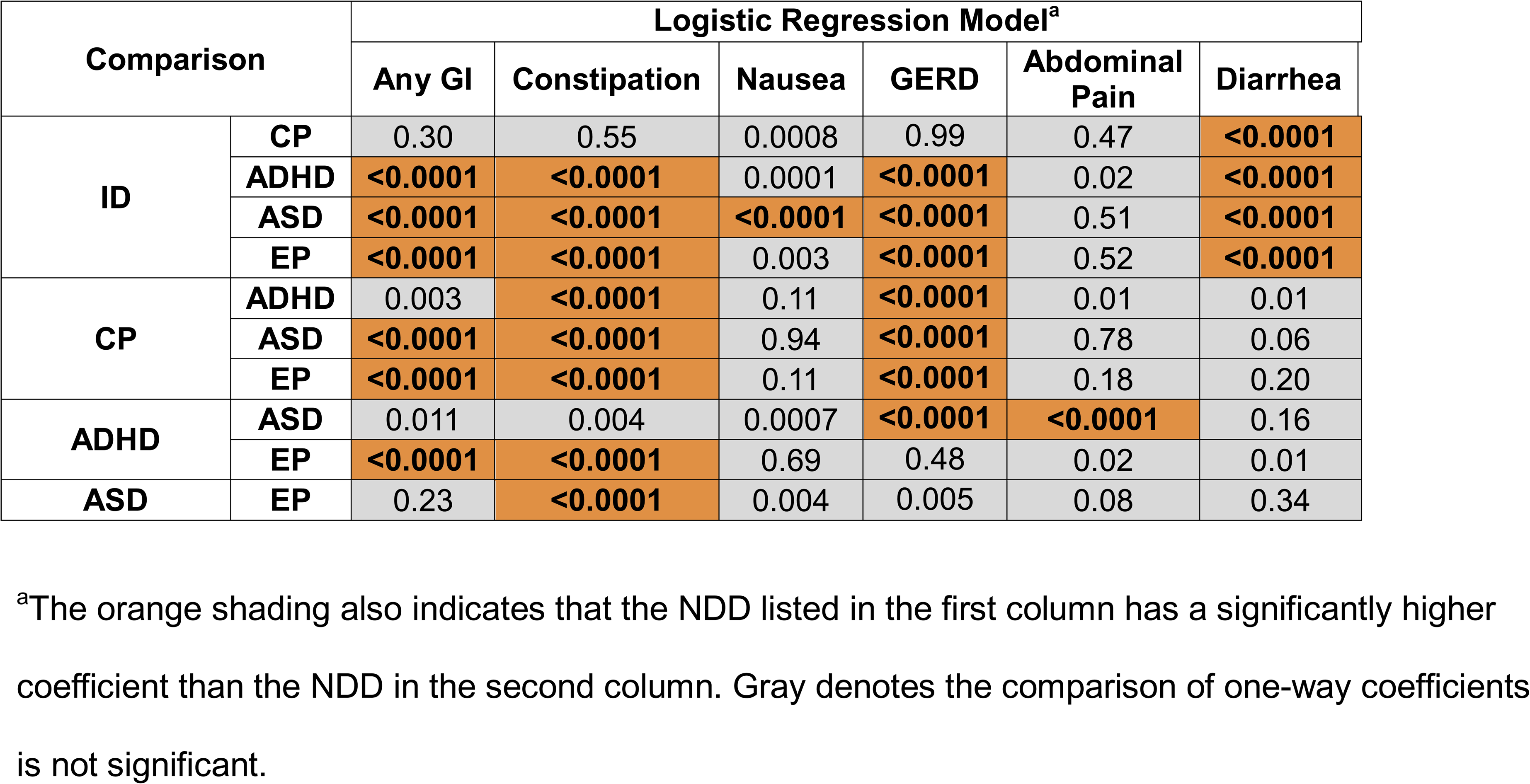
Hypothesis test comparisons between NDD coefficients in logistic regression models.

| Comparison |  | Logistic Regression Model<sup>a</sup> |  |  |  |  |  |
| --- | --- | --- | --- | --- | --- | --- | --- |
|  |  | Any GI | Constipation | Nausea | GERD | Abdominal Pain | Diarrhea |
| ID | CP | 0.30 | 0.55 | 0.0008 | 0.99 | 0.47 | <0.0001 |
|  | ADHD | <0.0001 | <0.0001 | 0.0001 | <0.0001 | 0.02 | <0.0001 |
|  | ASD | <0.0001 | <0.0001 | <0.0001 | <0.0001 | 0.51 | <0.0001 |
|  | EP | <0.0001 | <0.0001 | 0.003 | <0.0001 | 0.52 | <0.0001 |
| CP | ADHD | 0.003 | <0.0001 | 0.11 | <0.0001 | 0.01 | 0.01 |
|  | ASD | <0.0001 | <0.0001 | 0.94 | <0.0001 | 0.78 | 0.06 |
|  | EP | <0.0001 | <0.0001 | 0.11 | <0.0001 | 0.18 | 0.20 |
| ADHD | ASD | 0.011 | 0.004 | 0.0007 | <0.0001 | <0.0001 | 0.16 |
|  | EP | <0.0001 | <0.0001 | 0.69 | 0.48 | 0.02 | 0.01 |
| ASD | EP | 0.23 | <0.0001 | 0.004 | 0.005 | 0.08 | 0.34 |

<sup>a</sup>The orange shading also indicates that the NDD listed in the first column has a significantly higher coefficient than the NDD in the second column. Gray denotes the comparison of one-way coefficients is not significant.

## Discussion

By leveraging a large pediatric cohort of 42,204 individuals with matched controls, examining aggregate and condition-specific GI outcomes and quantifying associations across individual NDDs within a single health system, this study provides the largest and most comprehensive assessment of clinical GI morbidity across pediatric NDDs to date. The study demonstrates that all NDDs examined were associated with elevated rates of GI conditions relative to controls. The scale and design of this study strengthen its contribution to the literature that has often been limited by failure to account for coexisting NDDs, variable ascertainment of NDDs and GI conditions, and inconsistent findings regarding disorder-specific associations. The study population is larger than the combined size of prior studies directly comparing GI outcomes across ASD, other NDDs, and control groups^24–26,36–40^, and the use of a unified EHR system allowed GI diagnoses to be assessed consistently across all groups.

In this large pediatric EHR cohort, univariate analyses showed aggregate GI conditions were significantly more common among children with an NDD compared to matched controls. Having an NDD was also associated with significantly higher frequency of feeding difficulties and 10 of 14 individual GI conditions studied. Many elevations persisted after restricting the analysis to children with a single NDD, indicating that the increased GI burden was not explained solely by having multiple co-occurring NDDs. The association between NDDs and the presence of any GI condition was distributed across diagnoses and was not driven by one or two individual NDDs or a small number of individual GI conditions. For each of the five NDDs studied, cases had a significantly higher likelihood of having a GI condition compared to matched controls. Furthermore, each NDD was associated with statistically increased frequency of at least half (range 7-9) of the 14 individual GI conditions studied and with feeding difficulties, showing the association with any GI condition was not driven by a small number of individual GI conditions. Although statistical significance was found in many instances, the range of differences varied from small to substantial.

To facilitate comparison of NDD-GI associations among individual NDDs and identify potential diagnosis-specific relationships, effect sizes (ORs) were estimated for each NDD across aggregate GI conditions and the five most common individual GI conditions using multivariable models that simultaneously accounted for comorbid NDDs. Consistent with the univariate analyses, the odds of having any GI condition were significantly greater than 1.0 for each of the five NDDs. These findings provide evidence that GI morbidity is broadly elevated across NDDs even after accounting for diagnostic co-occurrence, highlighting the importance of transdiagnostic perspectives in both research and clinical care.

Pairwise comparisons of one-way coefficients among NDDs showed that ID and CP were most strongly associated with having any GI condition, constipation, and GERD. ID was most strongly associated with diarrhea. Notably, ASD was not unique among NDDs in its association with GI dysfunction, and it was not the among the NDDs most strongly associated with aggregate GI problems or any of the five GI conditions examined in the regression analyses. These results demonstrate that GI conditions are significantly more common among children with ASD than among controls without an NDD but are at odds with previous studies that found significantly more GI problems in children with ASD compared to children with other NDDs^25,26,36–38^.

The finding that aggregate GI problems are significantly more common among children with NDDs compared to controls is consistent with previous studies, and the most common GI conditions involved are well-aligned^3,7,12,14,15,19,24^. For example, in the present study, CP and ID were strongly associated with constipation, as reported in prior literature^14,15,29^, and the 33% frequency of GI conditions or feeding difficulty among children with ASD is within the 33-49% range reported by recent systematic reviews and meta-analyses^14,15,29^. Some previously reported associations, such as that between ADHD and irritable bowel syndrome^13^, were not confirmed^14,15,29^. Additionally, relationships between ASD and Crohn’s disease and ulcerative colitis were not observed, though this could be due to low counts across cases and controls, differences in ages of populations assessed, and the approach to capturing diagnoses^55^.

GI conditions are complex and may result from multiple etiologies simultaneously. Developmental and behavioral differences (e.g., stool withholding, restrictive diets, and anxiety), neurologic abnormalities (e.g., altered muscle tone and autonomic dysfunction), medication and supplement exposure, immunological and/or microbiome differences, and underlying genetic etiologies that affect the enteric nervous system may all contribute to the association between NDDs and GI dysfunction^26,56,57^. A growing body of evidence suggests that variants in high-confidence ASD genes (which are also high-confidence ID genes) affect both brain and ENS development^56–58^. Such variants lead to disruption of embryonic enteric neuronal progenitor cell migration, causing intestinal dysmotility and associated GI conditions such as constipation^56^. Given the overlap in potential contributing behaviors and neurologic abnormalities, shared genetic etiologies across NDDs^56,59–61^, and absence of evidence of GI pathology specific to ASD^31,32^, it is, perhaps, not surprising the study did not find evidence of a unique relationship between ASD and GI conditions.

Among children with NDDs, GI conditions are associated with substantial discomfort, functional limitations, severe disruptive behaviors, psychiatric comorbidities, and increased healthcare utilization^21,23–27,62^. Clinician awareness of GI comorbidity across NDDs may facilitate identification and initiation of appropriate treatment of these conditions. Documentation of GI and other comorbidity patterns across NDDs may help to identify subgroups relevant to treatment response and may provide insight into etiologic and phenotypic heterogeneity of NDDs^63^.

### Limitations

Inclusion criteria and study design were carefully selected to minimize potential limitations inherent to retrospective EHR-based ascertainment, yet limitations remain. The study population came from a single, relatively homogeneous health system and should be replicated in broader, more diverse cohorts. Additionally, reliance on EHR diagnostic codes to ascertain diagnoses may lead to classification errors; the absence of a diagnosis does not necessarily mean the absence of a condition (e.g., due to outside care, not seeking care for certain conditions, communication difficulties limiting symptom reporting). Further, cases and controls who interacted with the system more frequently may have had more opportunity for diagnostic capture. However, despite these limitations, the frequencies of the five NDDs studied were very similar to United States prevalence estimates from national survey data^1,2^. This study did not estimate condition severity and did not incorporate other variables that could impact GI conditions such psychotropic medication use. Furthermore, GI diagnoses were identified via a single set of criteria for all conditions based on diagnosis codes, and some GI diagnoses may represent more chronic concerns while others are indicative of illness. Future work should examine relevant quantitative traits and measures of severity and evaluate the association of GI dysfunction with psychiatric comorbidities, healthcare utilization, and other resource use. Finally, genetic factors were not evaluated in this study but should be incorporated in future investigations to clarify the contributions of large-effect, rare variants and polygenic risk to GI dysfunction across NDDs.

### Conclusions

Clinically recognized GI morbidity is elevated across a range of NDDs in children, and study findings do not provide evidence of a unique relationship between ASD and GI dysfunction. Although previous work has led to prioritization of ASD over the broader group of NDDs in research, management guidelines, and development of clinical tools, our findings do not support this emphasis and, instead, suggest broader efforts across NDDs are warranted.

## Supporting information

eTables 1-8

eTable 9

## Data Availability

Datasets generated during and/or analyzed during the current study are not publicly available, but aggregate data is obtainable from the corresponding author upon reasonable request.

## Code Availability

All code used in analyses is available at https://github.com/michellepistner/ndd_gi.

## Author Contributions

Concept and design: AJ, JMS, MPN, SMM, TDC

Acquisition, analysis, or interpretation of data: AJ, ASFB, CLM, DHL, JMS, LKW, MPN, SMM

Drafting of the manuscript: ASFB, JMS, MPN, SMM

Critical review of the manuscript for important intellectual content: AJ, ASFB, CLM, DHL, JMS, LKW, MPN, SMM, TDC

Statistical analysis: AJ, MPN

Administrative, technical, or material support:

N/A Supervision: CLM, DHL, SMM, TDC

## Acknowledgments

Research reported in this publication is supported by the National Institute of Mental Health (NIMH) of the National Institutes of Health (NIH) under award number R01MH074090 and the Eunice Kennedy Shriver National Institute of Child Health and Human Development (NICHD) of the NIH under award number R01HD104938.The funding bodies did not have a role in the design of the study or writing of the manuscript.

We acknowledge Geisinger patients who made this work possible. We would also like to acknowledge Celia Gray with the Geisinger Data Core for her guidance and support related to EHR data collection to support this study. We also would like to acknowledge Hannah Liefeld, MD for her efforts on an earlier version of this project and Elle King for her contributions to the literature review.

