## Supplementary material for "Elevated Rates of Gastrointestinal Dysfunction in Children with Neurodevelopmental Disabilities: Not Just an Autism Issue": eTables 1-8

### Supplemental Materials Table of Contents

### eTable 1. Coding Definitions for Considered NDD and GI Conditions

| Neurodevelopmental Disabilities Included in Study | | |
|---|---|---|
| **Diagnosis** | **ICD-9 Codes** | **ICD-10 Codes** |
| *Autism Spectrum Disorder (ASD)* | 299.00, 299.01, 299.10, 299.80, 299.81, 299.90 | F84, F84. * |
| *Intellectual Disabilities (ID)* | 317, 318.0, 318.1, 318.2, 319 | F70, F71, F72, F73, F78, F79 |
| Attention-deficit/hyperactivity disorder *(ADHD)* | 314, 314.0, 314.00, 314.01, 314.1, 314.2, 314.8, 314.9 | F90, F90.0, F90.1, F90.2, F90.8, F90.9 |
| *Epilepsy (EP)* | 345, 345. *, 345.0*, 345.1*, 345.4*, 345.5*, 346.6*, 345.7*, 345.8*, 345.9* | G40, G40. *, G40. **. G40. ***, G40.A, G40.A*, G40.A**, G40.B, G40.B*, G40.B** |
| *Cerebral Palsy (CP)* | 343, 343. * | G80, G80. * |
| **Neurodevelopmental Disabilities Considered but Not Included in Study** | | |
| **Diagnosis** | **ICD-9 Codes** | **ICD-10 Codes** |
| *Communication Disorders* | 315.3, 315.31, 315.32, 315.34, 315.35, 315.39, 307.0, 307.9 | F80.0, F80.1, F80.2, F80.4, F80.8, F80.81, F80.82, F80.89, F80.9, F98.5 |
| *Specific Learning Disorders* | 315.0, 315.0*, 315.1, 315.2 | F81, F81.0, F81.2, F81.8, F81.81, F81.89, F81.9 |
| *DSM-5 Motor Disorders* | 315.4, 307.2, 307.20, 307.21, 307.22, 307.23, 307.3, 781.3 | F82, F98.4, F95, F95.0, F95.1, F95.2, F95.8, F95.9, R27.0, R27.8, R27.9 |
| *Other Neurodevelopmental Disorders* | 315.9, 315.8, 315.5 | F89, F88 |
| **Gastrointestinal Disorders** | | |
| **Diagnosis** | **ICD-9** | **ICD-10** |
| *Constipation* | 564.0* | K59.0, K59.0* |
| *Diarrhea* | 564.5, 787.91 | K59.1, R19.7 |
| *Abdominal pain/tenderness* | 789.00, 789.01, 789.02, 789.03, 789.04, 789.05, 789.07, 789.09, 789.6*, | R10.10, R10.11, R10.12, R10.3, R10.3*, R10.8*, R10.81*, R10.82*, R10.9 |
| *Functional dyspepsia (indigestion), heartburn* | 536.8, 787.1, 789.06 | K30, R12, R10.13 |
| *Nausea and vomiting* | 536.2, 569.87, 787.0* | R11. *, R11.1* |
| *Flatulence and related conditions* | 787.3 | R14. * |
| *Irritable bowel syndrome (IBS)* | 564.1 | K58.*R |
| *Gastroesophageal reflux disease (GERD)* | 530.81, 530.11, 530.81 | K21.*, K21.0* |
| *Eosinophilic esophagitis (EoE)* | 530.13 | K20.0 |
| *Gastroparesis* | | K31.84 |

| | | |
|---|---|---|
| *Crohn's disease (CD)* | 555.* | K50.*, K50.0*, K50.01*, K50.1*, K50.11*, K50.8*, K50.81*, K50.9*, K50.91* |
| *Ulcerative colitis (UC)* | 556.* | K51.*, K51.0*, K51.01*, K51.2*, K51.21*, K51.3*, K51.31*, K51.4*, K51.41*, K51.5*, K51.51*, K51.8*, K51.81*, K51.9*, K51.91* |
| *Other noninfectious colitis* | 558.3, 558.41, 558.9, 535.70, 535.71 | K52.2*, K52.3, K52.8*, K52.83*, K52.89, K52.9 |
| *Intestinal malabsorption* | 579.* | K90.* |
| *Feeding difficulties* | 783.3 | R63.3* |

Abbreviations: DSM-5, Diagnostic and Statistical Manual of Mental Disorders, Fifth Edition; ICD, International Classification of Diseases; GI, gastrointestinal; NDD, neurodevelopmental disabilities

* Denotes that all child codes of that parent code are included

### eTable 2. Counts of Neurodevelopmental Disability (NDD) Diagnoses

| | **Number with this NDD diagnosis** | **Number with *only* this NDD diagnosis** |
|---|---|---|
| *Any NDD*[a] | 42,204 | NA |
| *ADHD* | 31,832 | 25,200 |
| *ASD* | 9,241 | 3,219 |
| *CP* | 1,645 | 633 |
| *EP* | 5,761 | 3,097 |
| *ID* | 5,173 | 1,045 |

Abbreviations: ADHD, attention-deficit/hyperactivity disorder; ASD, autism spectrum disorder; CP, cerebral palsy; EP, epilepsy; ID, Intellectual disabilities; NDD, neurodevelopmental disability
[a] Any NDD is defined as a diagnosis of attention-deficit/hyperactivity disorder (ADHD), autism spectrum disorder (ASD), cerebral palsy (CP), epilepsy (EP), or intellectual disabilities (ID)

### eTable 3. Frequency of Studied Neurodevelopmental Diagnoses in Geisinger Cohort and National Health Interview Surveys

| | Geisinger Pediatric Cohort (1989-2021, n=339,606, age 0-17)[a] | National Health Interview Survey (2018-2021, n=26,422, age 3-17)[1] | National Health Interview Survey (2009-2017, n=88,530, age 3-17)[2] |
|---|---|---|---|
| ADHD | 9.4% *(n=31,832)* | 9.57% *(n=2,679)* | 9.04% *(n=7,918)* |
| ASD | 2.7% *(n=9,241)* | 2.94% *(n=811)* | 1.74% *(n=1,550)* |
| CP | 0.5% *(n=1,645)* | NA | 0.31% *(n=264)* |
| EP | 1.7% *(n=5,761)* | NA | 0.77%[b] *(n=668)* |
| ID | 1.5% *(n=5,173)* | 1.72% *(n=446)* | 1.10% *(n=1,021)* |

Abbreviations: ADHD, attention-deficit/hyperactivity disorder; ASD, autism spectrum disorder; CP, cerebral palsy; EP, epilepsy; ID, Intellectual disabilities; NDD, neurodevelopmental disability

[a] Includes all patients meeting birthday criteria, with ≥6 months of electronic health record data, known sex. Excludes patients with only one of the four excluded NDDs – communication disorders, DSM-5 motor disorders, specific learning disorders, and other neurodevelopmental disorders.

[b]Defined as seizure in the last 12 months in the National Health Interview Survey

### eTable 4. NDD Case Demographic Comparison by Number of Conditions

| | **One NDD Condition** N = 33,194 | **Two Plus NDD Conditions** N = 9,010 | **p-value**[a] |
|---|---|---|---|
| Patient sex | | | <0.0001 |
| Female | 10,682 (32.18%) | 2,445 (27.14%) | |
| Male | 22,512 (67.82%) | 6,565 (72.86%) | |
| Patient race | | | <0.0001 |
| Black or African American | 2,316 (6.98%) | 711 (7.89%) | |
| Other | 451 (1.36%) | 167 (1.85%) | |
| Unknown | 262 (0.79%) | 78 (0.87%) | |
| White | 30,165 (90.87%) | 8,054 (89.39%) | |
| Age at Last Encounter | 15.13 (11.01, 17.53) | 14.04 (9.68, 17.39) | <0.0001 |
| Length of EHR (Years) | 7.70 (4.25, 11.99) | 8.03 (4.68, 12.23) | <0.0001 |
| Has any GI condition | 9,826 (29.60%) | 3,284 (36.45%) | <0.0001 |
| Has any GI condition **or** Feeding Difficulty | 10,064 (30.32%) | 3,489 (38.72%) | <0.0001 |

Abbreviations: EHR, electronic health record; GI, gastrointestinal; NDD, neurodevelopmental disability

[a]Pearson's Chi-squared test; Wilcoxon rank sum test. For all tests, a Bonferroni-adjusted α of 0.0001 was used

### eTable 5. Demographic Variables Compared Between NDD Cases and Controls

| | **NDD Case** N = 42,204 n (%); Median (Q1, Q3) | **NDD Control** N = 297,402 n (%); Median (Q1, Q3) | **p-value**[a] |
|---|---|---|---|
| Patient Sex | | | <0.0001 |
| Female | 13,127 (31.10%) | 154,443 (51.93%) | |
| Male | 29,077 (68.90%) | 142,959 (48.07%) | |
| Patient race | | | <0.0001 |
| Black or African American | 3,027 (7.17%) | 21,447 (7.21%) | |
| Other | 618 (1.46%) | 8,050 (2.71%) | |
| Unknown | 340 (0.81%) | 3,014 (1.01%) | |
| White | 38,219 (90.56%) | 264,891 (89.07%) | |
| Age at Last Encounter in Study Period | 14.92 (10.69, 17.51) | 13.89 (8.23, 17.28) | <0.0001 |
| Length of EHR (Years) | 7.76 (4.34, 12.03) | 5.60 (2.63, 9.73) | <0.0001 |
| Has any GI condition | 13, 110 (31.06%) | 60, 246 (20.26%) | <0.0001 |
| Has any GI condition **or** Feeding Difficulty | 13,553 (32.11%) | 61,168 (20.57%) | <0.0001 |

Abbreviations: EHR, electronic health record; GI, gastrointestinal; NDD, neurodevelopmental disability

[a] Pearson's Chi-squared test; Wilcoxon rank sum test. For all tests, a Bonferroni-adjusted α of 0.0001 was used.

### eTable 6. Frequency of Various GI Diagnoses on Unmatched NDD Cases and Controls.

| **GI Diagnosis** | **NDD Case** N = 42,204 n (%) | **Control** N = 297,402 n (%) | **p-value**[a] |
|---|---|---|---|
| Constipation | 5,415 (12.83%) | 17,432 (5.86%) | <0.0001 |
| Diarrhea | 2,875 (6.81%) | 10,508 (3.53%) | <0.0001 |
| Abdominal pain or tenderness | 3,876 (9.18%) | 20,956 (7.05%) | <0.0001 |
| Functional dyspepsia (indigestion) or heartburn | 863 (2.04%) | 4,380 (1.47%) | <0.0001 |
| Nausea or vomiting | 4,286 (10.16%) | 17,253 (5.80%) | <0.0001 |
| Flatulence and related conditions | 271 (0.64%) | 1,022 (0.34%) | <0.0001 |
| Irritable bowel syndrome (IBS) | 264 (0.63%) | 1,716 (0.58%) | 0.22 |
| Gastroesophageal reflux disease (GERD) | 5,200 (12.32%) | 20,305 (6.83%) | <0.0001 |
| Eosinophilic esophagitis (EoE) | 151 (0.36%) | 551 (0.19%) | <0.0001 |
| Gastroparesis | 199 (0.47%) | 481 (0.16%) | <0.0001 |
| Crohn's Disease (CD) | 85 (0.20%) | 569 (0.19%) | 0.66 |
| Ulcerative colitis (UC) | 48 (0.11%) | 285 (0.10%) | 0.27 |
| Other noninfectious colitis | 1,041 (2.47%) | 4,221 (1.42%) | <0.0001 |
| Intestinal malabsorption | 393 (0.93%) | 1,830 (0.62%) | <0.0001 |
| Any GI diagnosis | 13,110 (31.06%) | 60,246 (20.26%) | <0.0001 |
| Feeding difficulties | 1,855 (4.40%) | 2,339 (0.79%) | <0.0001 |
| Any GI condition **or** Feeding difficulties | 13,553 (32.11%) | 61,168 (20.57%) | <0.0001 |

Abbreviations: GI, gastrointestinal; NDD, neurodevelopmental disability
[a]Pearson's Chi-squared test. For all tests, a Bonferroni-adjusted α of 0.0001 was used.

### eTable 7. Demographic Comparison of Matched Cohort

| | **NDD Case** N = 42,204 n (%); Median (Q1, Q3) | **Control** N = 84,405 n (%); Median (Q1, Q3) | **p-value**[a] |
|---|---|---|---|
| Patient sex | | | >0.9 |
| Female | 13,127 (31.10%) | 26,253 (31.10%) | |
| Male | 29,077 (68.90%) | 58,152 (68.90%) | |
| Patient race | | | >0.9 |
| Black or African American | 3,027 (7.17%) | 6,054 (7.17%) | |
| Other | 618 (1.46%) | 1,236 (1.46%) | |
| Unknown | 340 (0.81%) | 677 (0.80%) | |
| White | 38,219 (90.56%) | 76,438 (90.56%) | |
| Age at Last Encounter | 14.92 (10.69, 17.51) | 15.01 (10.69, 17.44) | 0.07 |
| Length of EHR (Years) | 7.76 (4.34, 12.03) | 7.77 (4.34, 12.05) | 0.73 |

Abbreviations: EHR, electronic health record; NDD, neurodevelopmental disability
[a]Pearson's Chi-squared test; Wilcoxon rank sum test. For all tests, a Bonferroni-adjusted α of 0.0001 was used.

### eTable 8. Frequency of GI Condition by NDD

| | Any GI Condition | | | Any GI Condition *or* Feeding Difficulty | | |
|---|---|---|---|---|---|---|
| | **Any GI Condition** | **Any GI Condition in Matched Controls** | **p-value[a]** | **Any Condition or Feeding Difficulty** | **Any GI Condition or Feeding Difficulty in Matched Controls** | **p-value[a]** |
| *Any NDD (N=42,204)* | 13,110 (31.06%) | 18,816 (22.29%) | <0.0001 | 13,553 (32.11%) | 19,052 (22.57%) | <0.0001 |
| *ADHD (N=31,832)* | 9,699 (30.47%) | 14,302 (22.47%) | <0.0001 | 9,907 (31.12%) | 14,467 (22.72%) | <0.0001 |
| *ASD (N = 9,241)* | 2,861 (30.96%) | 4,034 (21.83%) | <0.0001 | 3,072 (33.24%) | 4,088 (22.12%) | <0.0001 |
| *CP (N = 1,645)* | 815 (49.54%) | 735 (22.35%) | <0.0001 | 859 (52.22%) | 745 (22.65%) | <0.0001 |
| *EP (N = 5,761)* | 2,030 (35.24%) | 2,684 (23.29%) | <0.0001 | 2,103 (36.50%) | 2,720 (23.61%) | <0.0001 |
| *ID (N = 5,173)* | 2,144 (41.45%) | 2,456 (23.74%) | <0.0001 | 2,334 (45.12%) | 2,502 (24.19%) | <0.0001 |

Abbreviations: ADHD, attention-deficit/hyperactivity disorder; ASD, autism spectrum disorder; CP, cerebral palsy; EP, epilepsy; GI, gastrointestinal; ID, Intellectual disabilities; NDD, neurodevelopmental disability

[a] For all tests, a Bonferroni-adjusted α of 0.0001 was used.

1 Li, Q. *et al.* Prevalence and trends of developmental disabilities among US children and adolescents aged 3 to 17 years, 2018-2021. *Sci Rep* **13**, 17254 (2023). https://doi.org/10.1038/s41598-023-44472-1

2 Zablotsky, B. *et al.* Prevalence and Trends of Developmental Disabilities among Children in the United States: 2009-2017. *Pediatrics* **144** (2019). https://doi.org/10.1542/peds.2019-0811
